# Spotlight on the dark figure: Exhibiting dynamics in the case detection ratio of COVID-19 infections in Germany

**DOI:** 10.1101/2020.12.23.20248763

**Authors:** Marc Schneble, Giacomo De Nicola, Göran Kauermann, Ursula Berger

## Abstract

The case detection ratio of COVID-19 infections varies over time due to changing testing capacities, modified testing strategies and also, apparently, due to the dynamics in the number of infected itself. In this paper we investigate these dynamics by jointly looking at the reported number of detected COVID-19 infections with non-fatal and fatal outcomes in different age groups in Germany. We propose a statistical approach that allows us to spotlight the case detection ratio and quantify its changes over time. With this we can adjust the case counts reported at different time points so that they become comparable. Moreover we can explore the temporal development of the real number of infections, shedding light on the dark number. The results show that the case detection ratio has increased and, depending on the age group, is four to six times higher at the beginning of the second wave compared to what it was at the peak of the first wave. The true number of infection in Germany in October was considerably lower as during the peak of the first wave, where only a small fraction of COVID-19 infections were detected. Our modelling approach also allows quantifying the effects of different testing strategies on the case detection ratio. The analysis of the dynamics in the case detection rate and in the true infection figures enables a clearer picture of the course of the COVID-19 pandemic.

## 1. Introduction

Originating from Wuhan, China, COVID-19 developed to become a worldwide pandemic in the Spring of 2020 (Velavan and Meyer, 2020). Starting from the very beginning of this unprecedented health crisis, the issue of case detection, while always being at the center of scientific and public discourse, has been all but transparent. Knowing how many infections are really present in the population would be of paramount importance, and researchers have tried to tackle the problem in several different ways. Early in the epidemic wave, the ratio of undetected COVID-19 cases was likely to be high, i.e. 5 to 20 times higher than the number of confirmed cases (e.g. Li et al., 2020 or Wu et al., 2020). The problem of discovering the case detection ratio is tightly intertwined with the uncovering of the true fatality ratio of the disease, as knowledge on one of those two unknown quantities would provide a large amount of information about the other. A natural experiment which allowed to estimate both the fatality ratio and the case detection ratio occurred with the outbreak on the cruise ship “Diamond Princess” (Mizumoto et al., 2020). During the early stages of the pandemic, the true percentage of the population infected for eleven European countries was deduced from early estimates of the mortality rates (Flaxman et al., 2020). Moreover, Aspelund et al. (2020) used Bayes arguments applied to testing data from Ireland to estimate the case detection ratio in the order of 7-11% in the beginning of the pandemic and in the order of 10-20% thereafter. The argument is based on relating the number of tests and the share of positive tests. A similar approach has been pursued making use of Canadian data (Benatia et al., 2020). The problem of estimating the true numbers of COVID-19 infections has also been discussed from a purely statistical point of view, where the case detection ratio was related to the fatality ratio (Manski and Molinari, 2020). Overall, underreporting appears to be an overarching problem, which plays a central role when estimating the case detection ratio for COVID-19 (Russell et al., 2020). The importance to assess the detection ratio and its effect on predictions of the pandemic’s course has been demonstrated in mathematical simulation studies (Fuhrmann and Barbarossa, 2020). In this context, different national underreporting ratios have been compared (e.g. Rahmandad et al., 2020 or Jagodnik et al., 2020) and a general discussion and survey on assessing the infection fatality ratio was conducted (Levin et al., 2020).

It is clear that generally the case detection ratio changes greatly over time depending on testing capacities and different testing strategies being employed, that vary with time and region. In Germany the number of test has increased considerably since the outbreak of the pandemic in March 2020. Also the testing strategy has been adjusted over the course of the pandemic, where in the beginning mainly individuals with symptoms have been tested, in later phases a enormous number of tests as been performed on travellers returning from foreign countries and contact persons of COVID-19 positive individuals. In this paper, we explore the dynamics in the case detection ratio using the number of infections in Germany from March to October 2020. Thereto we propose an approach that relates fatal cases of COVID-19 to non-fatal ones, detected through testing. The latter are either cured cases or infections without any symptoms. We make use of publicly available registry data of COVID-19 cases provided by the Robert-Koch-Institute (Esri Deutschland GmbH, 2020). While with the given data a quantification of the case detection ratio is not possible in absolute terms, we can derive the case detection ratio on a relative scale, that is we can show how the case detection ratio changed over the course of the pandemic. The statistical idea behind this leads to a dynamic generalized linear mixed model with smooth random effects (see e.g. Durbán et al., 2005, Durban and Aguilera-Morillo, 2017 and Wood, 2017). The major advantage of our approach is that it only relies on the assumption that the age specific COVID-19 fatality ratio, whatever it is, has not substantial changed over time. Whether this assumption is valid is currently discussed (Harris, 2020; Kip et al., 2020) and the possibility of changed fatality ratios in the second wave has been considered as well (Kenyon, 2020 and Aspelund et al., 2020). To assess the impact of this assumption on our results, we provide sensitivity analyses that demonstrate that our approach is sufficiently robust.

Our focus is on exhibiting and evaluating the temporal development in the case detection ratio, which allows us to shed light on the dark number of COVID-19 infections. With it, we explore the dynamics in the real numbers of infections. In our analysis, we look at data from Germany and fit separate models for different age groups. We also carry out our analysis on a finer resolution, breaking it down to the federal state level in Germany. The latter allows us to evaluate the effect of different testing strategies pursued in different regions.

## 2. Data

We make use of COVID-19 data openly provided by the Robert-Koch-Institute (RKI), the German federal government agency and scientific institute responsible for health reporting, disease control and prevention in humans (Esri Deutschland GmbH, 2020). The data, exemplified in Table 1, contain cumulated counts of newly registered, laboratory-confirmed COVID-19 cases in Germany for each calendar day stratified by age group (0-4, 5-14, 15-34, 35-59, 60-79 or 80+ years), gender (male/female) and district (412 in total). Furthermore, for all registration dates and strata, the number of deaths associated with COVID-19 transmitted to the RKI by the local health authorities of the respective district is recorded. Note that the date of death is not provided, but for each death we have the date when the infection was detected by a test. The database of the Robert-Koch-Institute is updated every morning with the new numbers transmitted to it from the local health authorities.

**Table 1:** Illustration of the data structure. To facilitate reproducibility, the original column names used in the RKI dataset are given in brackets below our English notation.

| District<br>(Landkreis) | Age Group<br>(Altersgruppe) | Gender<br>(Geschlecht) | Cases<br>(Anzahl Fall) | Deaths<br>(Anzahl Todesfall) | Registration Date<br>(Meldedatum) |
| --- | --- | --- | --- | --- | --- |
| ⋮ | ⋮ | ⋮ | ⋮ | ⋮ | ⋮ |
| Munich City | 60-79 | F | 26 | 0 | September 8, 2020 |
| Munich City | 60-79 | M | 21 | 1 | September 8, 2020 |
| ⋮ | ⋮ | ⋮ | ⋮ | ⋮ | ⋮ |

In this study, we only consider data entries with registration dates ranging from calendar week 10 to calendar week 47, since for earlier weeks the number of tests being positive was not large enough to draw conclusive results. On the other hand, we have knowledge on the empirical time span observed from registration as infected with COVID-19 to a fatal outcome reported by the RKI. The 95% quantile of this quantity is around 50 days. Therefore, by excluding registration dates up to seven weeks in the past from today, we ensure that the vast majority of all reported infections have either come to a fatal end or have recovered. In other words, for (almost) all cases included in our analysis we know the final outcome of the disease.

Moreover, although the data are given on a daily resolution, we here aggregate it into weekly data, which renders reporting delays occurring over the weekends and weekly reporting cycles irrelevant to our analysis, leading to more stable results. Since for children aged 14 years and younger barely any fatalities have been recorded, we excluded these age groups from our analysis.

## 3. Methods

When describing the dynamics of the COVID-19 pandemic, the number of interest is the true count of infected persons in a cohort, which shall be denoted by *I*_*t*_ for week *t* = 1, …, *T*. Note that *I*_*t*_ remains unobservable. The number can however be decomposed into the number of detected and reported cases *D*_*t*_ and the unknown number of infected persons, who have not been tested and remain undetected, i.e. the dark number *U*_*t*_. Hence, we have *I*_*t*_ = *D*_*t*_ + *U*_*t*_ and *D*_*t*_*/I*_*t*_ defines the case detection ratio, which however remains unknown due to the lack of knowledge of *U*_*t*_.

At timepoint *T* + 7(weeks) we have uncensored knowledge on the outcomes of all reported cases *D*_*t*_, that is, we know if they ended fatally or if they recovered. Consequently, the reported cases decompose into either recovered (non-fatal) outcomes *R*_*t*_ or fatal outcomes *F*_*t*_, i.e. *D*_*t*_ = *R*_*t*_ + *F*_*t*_. With this, the total number of infected persons splits into *I*_*t*_ = *R*_*t*_ + *F*_*t*_ + *U*_*t*_.

The expected number of reported fatal cases *F*_*t*_ as well as the expected number of recovered cases *R*_*t*_ are fractions of the total number of infections *I*_*t*_. This leads to

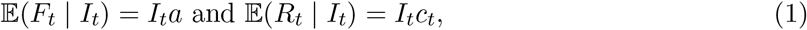

where 0 *<* (*a* + *c*_*t*_) *<* 1. Here, quantity *a* defines the Infection Fatality Ratio (IFR), while *c*_*t*_ is the Case Detection Ratio (CDR) of non-fatal (recovered) infections. Note that these non-fatal infections also include mild and symptom-free cases. Thus, if testing capacities are increased or the testing strategy is changed, *c*_*t*_ will change as well, which is incorporated in the notation by time index *t*. In contrast, the infection fatality ratio *a* will be assumed to remain constant over time. This can be justified by the fact that fatal cases, due to their severeness, are likely to be deteced independently of any testing policy. This also includes, to some extent, post-mortem tests.

With this notation we obtain the time-dependent case detection ratio CDR_*t*_ = *a* + *c*_*t*_. Note that for the dark number, i.e. the latent number of undetected infections *U*_*t*_, it holds that 𝔼(*U*_*t*_ | *I*_*t*_) = (1 − CDR_*t*_)*I*_*t*_. It would of course be favourable to estimate the number of undetected infections *U*_*t*_ via estimation of *a* and *c*_*t*_. However, when only the reported fatal and non-fatal cases *F*_*t*_ and *R*_*t*_ are known, these two ratios can not be estimated due to non-identifiability issues, which we will demonstrate below. Nonetheless, with the data at hand, we are able to estimate the ratio *c*_*t*_*/a*. To see this, we rewrite the above model in an equivalent form by defining a binary covariate *x* ∈ {0, 1} and by specifying the response variable *Y*_*t*_ through

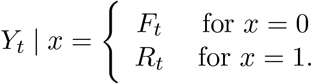

This notational trick allows us to rewrite the above relations (1) as a regression model

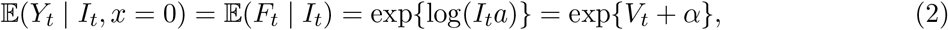

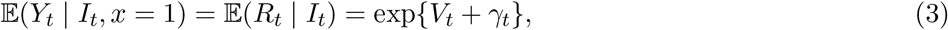

where *V*_*t*_ = log(*I*_*t*_), *α* = log(*a*) and *γ*_*t*_ = log(*c*_*t*_). Equations (2) and (3) can in turn be summarized into a single regression model formula

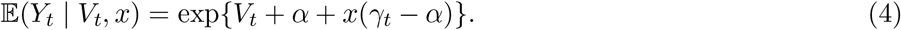

Note that *I*_*t*_ and hence *V*_*t*_ = log(*I*_*t*_) remain unobserved. We employ a Bayesian view and model *V*_*t*_ as normally distributed random effects *V*_*t*_ ∼ *N* (*µ*_*t*_, *σ*^2^). Still, the parameters in model (4) are not identifiable, since any shift in *µ*_*t*_ and a matching negative shift in *α* and *γ*_*t*_, respectively, results in the same model. This demonstrates the identifiability problem, which we have mentioned above. Hence, we are neither able to estimate the fatality ratio *a* = exp(*α*) nor the time-dependent ratio *c*_*t*_ = exp(*γ*_*t*_) with the data at hand. However, by setting *µ*_*t*_ = 0 and defining the global intercept *β*_0_ = *µ*_*t*_ + *α* we can rewrite model (4) to obtain the final regression model

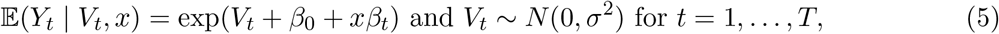

where *β*_*t*_ = *γ*_*t*_ − *α* and exp(*β*_*t*_) = *c*_*t*_*/a*. With this model we can now explore the dynamics in the the case detection ratio. For two different time points *t*_1_ and *t*_2_ we have

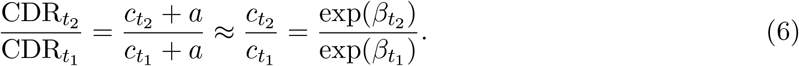

The approximation in (6) is justified by the fact that the number of fatal cases is much smaller than the number of recovered cases, i.e. *a ≪ c*_*t*_. Thus, 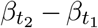 is a proxy for 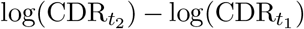 and exp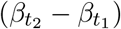 is a proxy for the relative change of the case detection ratio 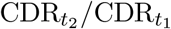.

The remaining question is how to model the dynamics in time *t*. It is natural to assume that changes in the case detection ratio over time do not occur suddenly, but gradually. For instance, test capacities are slowly increased and test strategies are gradually changed. To accommodate this in our model (5) we fit *β*_*t*_ by a smooth function in time leading to a time-varying coefficient model (Hastie and Tibshirani, 1993). We also induce smooth dynamics on the random component leading to a time-varying random effect (Durban and Aguilera-Morillo, 2017). These modifications lead to an identifiable and dynamic mixed regression model, for which we use a negative-binomial distribution for *Y*_*t*_ with a constant dispersion factor. The entire model can be fitted with standard software: all our analyses were performed in **R** (Team et al., 2013) and the dynamic mixed regression model is fitted using the **R**-package `mgcv` (Wood, 2017).

We apply this modelling approach using the reported data from calendar week 10 (beginning of March) to calendar week 47 (end of October), stratified by different age groups, to visualise the dynamics in the real infection numbers and the case detection ratio from the beginning of the pandemic up to the beginning of the second wave. To assess the robustness of the approach with respect to the assumption of time-constant and age-specific fatality ratios, we refit the model based on data excluding the time window of the first wave. The results for this are shown in the appendix.

Our main analyses are based upon data aggregated on the national level, while the original data are provided on the district level. For a regional view, we aggregate the data on the federal state level and regard the two federal states Bavaria and North-Rhine-Westphalia. These two federal states have reported the highest case numbers in Germany. The results for these regional analyses are also shown in the appendix.

## 4. Results

To give a first insight into the data at hand, we plot in Figure 1 the raw numbers of cases reported by the offical health authorities over time together with the raw number of fatalities stratified by age groups. This is shown in the top four plots on a log-scale. Both the number of registered cases and that of fatal cases (indexed by registration date of the infection, and not by day of death) peak in calendar week 13 for the two younger age groups and calendar week 14 for the two eldest age groups, respectively. Over the following weeks these numbers decrease. The small peak in calendar week 25 was caused by an outbreak in the district of Gütersloh, which is explained in more depth later. From calendar week 28 onward we resume to seeing an exponential increase of registered cases while the numbers of registered fatal cases only start to rise seven weeks later, also exponentially.

**Figure 1:**
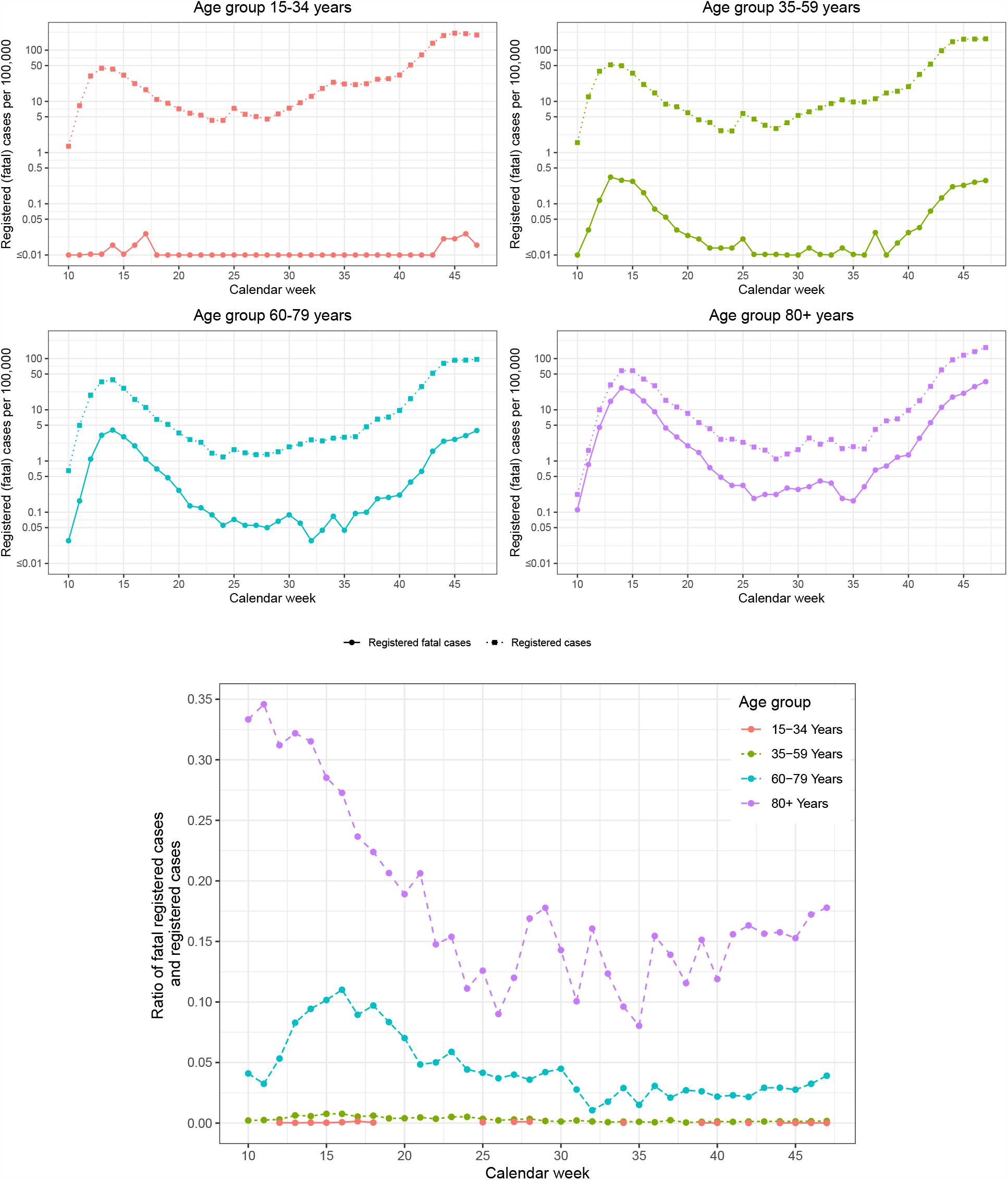
Raw data: registered cases of COVID-19 infections and registered fatal cases on a weekly basis for Germany. Top figure: Absolute numbers on a log-scale stratified by age group. Bottom figure: Case fatality ratios (= fatal cases / registered cases) stratified by age group.

The raw case fatality ratio, calculated as the ratio of fatal cases over total registered cases, stratified by age group, is shown in the bottom of Figure 1. The raw case fatality ratio in age group 80+ generally dropped from calendar week 10 onward and fluctuated mostly between 10% and 15% from week 25 onward. However, since calendar week 40 the case fatality ratio in this age group was throughout higher than 15%. In the age group 60-79 the case fatality ratio has peaked in calendar week 16 and gradually decreased to 2.5%. Here, we see as well a steady increase towards calendar week 47. All other age groups exhibit rather low raw case fatality ratios throughout.

Note that the raw data do not contain undetected cases (dark number) and therefore can not mirror the correct picture of the actual infection numbers, nor do these plots provide any information about the case detection ratio. In the following, we use the data to fit the statistical model described above, which enables us to estimate the relative change of the case detection ratio over time.

To explore the magnitude of undetected infections (i.e. the dark number), we fit the specified dynamic mixed model (5) with data from calendar weeks 10 - 47. Since the infection fatality ratio *a* depends on age, we fit separate models for each of the relevant age groups defined by the RKI, i.e. 15-34 years, 35-59 years, 60-79 years and 80+ years. The dynamics in the true infection numbers on the log-scale, represented by the fitted smooth dynamic random effects *V*_*t*_, are displayed in Figure 2. Note that these curves mirror the relative change over time, i.e. the dynamics, while the absolute numbers can not be interpreted on their own due to the mentioned identifiability problem. We therefore centered the curves around zero. We can see that the relative course of the pandemic was very similar across all age groups, where a peak is reached around calendar week (CW) 14. However, the peak for the younger age groups is estimated to be around one week earlier than for the older age groups, i.e. in CW 13. An explanation for this finding is that the younger age groups have been more affected by the lockdown, which started in Germany in calendar week 12. Further comparing the maximum max_*t*_ *V*_*t*_ and the minimum min_*t*_ *V*_*t*_ of *V*_*t*_, we see that the difference max_*t*_ *V*_*t*_ − min_*t*_ *V*_*t*_ increases with age, i.e. the relative decline in true infections numbers was less pronounced in the younger age groups. Also eye-catching is the increase in infections around calendar week 25 for people below 60 years of age. This is the aforementioned outbreak in the district of Gütersloh, which occurred in an industrial slaughterhouse and has mainly affected people in the working age. From CW 35 (end of August) all curves start rising steadily, where the steepest rise is seen for the oldest age group, while the rise is flatter for the younger the age group. This shows that the second wave of the pandemic had already begun around CW 35.

**Figure 2:**
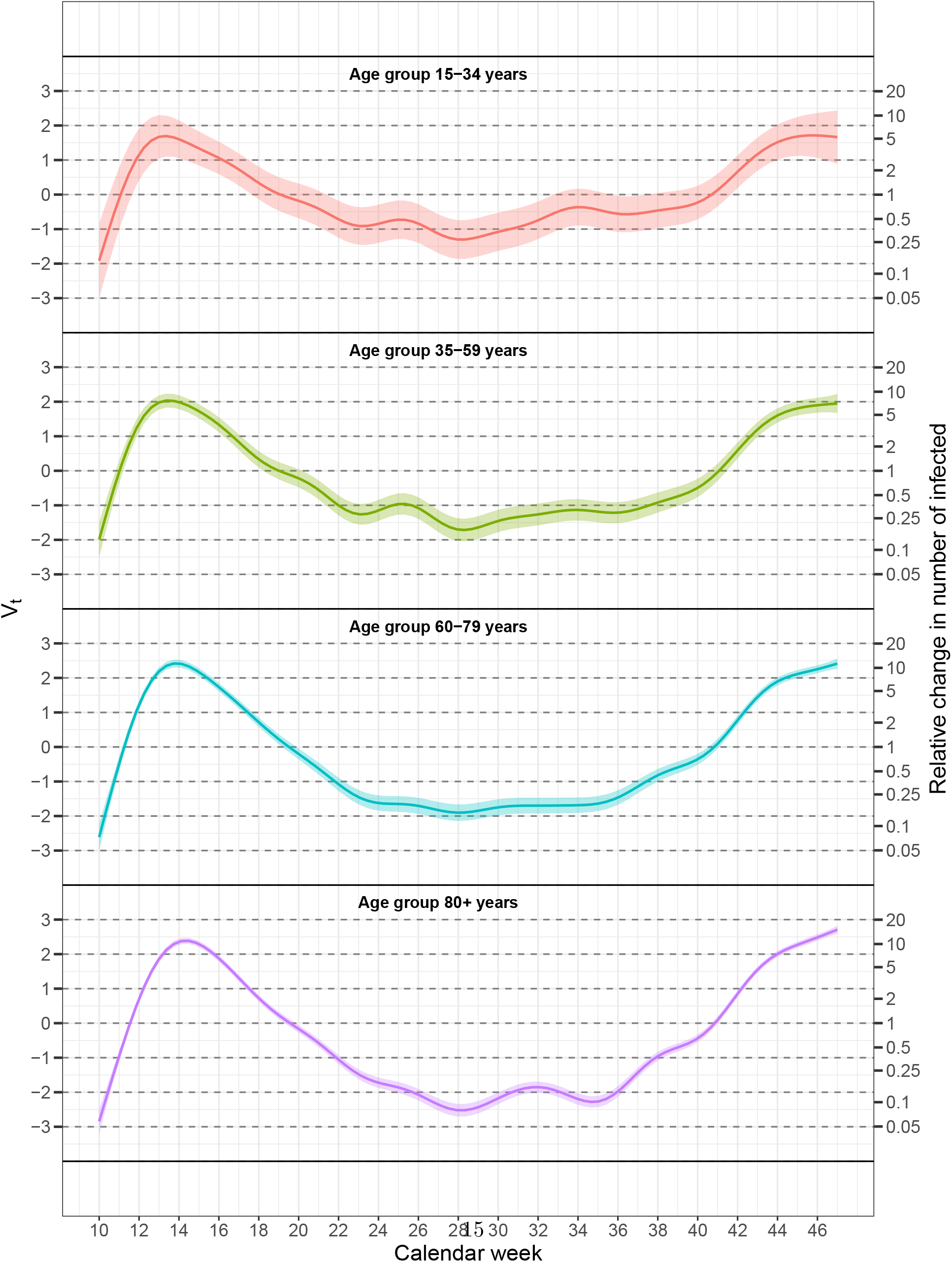
Dynamics of the true infection numbers on the log-scale for different age groups: The smooth random effects *V*_*t*_. The shaded areas represent 95% confidence bands.

Next, we look at the dynamics in the case detection ratio. Figure 3 shows the fitted time-varying coefficients *β*_*t*_ together with corresponding 95% confidence bands. Again, the absolute level is not identifiable, so these curves are normalized such that *β*_*CW* 10_ = 0. Hence, the function values on the exp-scale (right y-axes) give the relative change in the case detection ratio (CDR) with respect to calendar week 10. The case detection ratio in the age group 80+ has risen monotonically since the beginning of the pandemic up to calendar week 33, where our model estimates a more than four-times higher case detection ratio when compared to mid-March. Note that in later weeks, the case detection ratio among the elderly decreased again to the level of April/May. In contrast, for people aged 60 to 79, the case detection ratio first dropped by about 70%, reaching its bottom as the pandemic passed its peak in Germany in CW 16. We subsequently see a monotonic increase, with the case detection ratio becoming 1.5 times higher compared to the beginning of the pandemic. Again, this is followed by a slight decline towards CW 47. The dynamics in the case detection ratio in the population aged 35-59 years are similar to those of the 60 to 79 years old: after a drop during March and April (CW 10 - CW 16), CDR increases, in mid-September to nearly three times what it was in CW 10. For the youngest age group (aged 15-34) we also see a rise in the case detection ratio over time, which seems substantial. The confidence bands in this age group are, however, rather wide, due to the fact that this age group is not as prone to fatal outcomes as older age groups.

**Figure 3:**
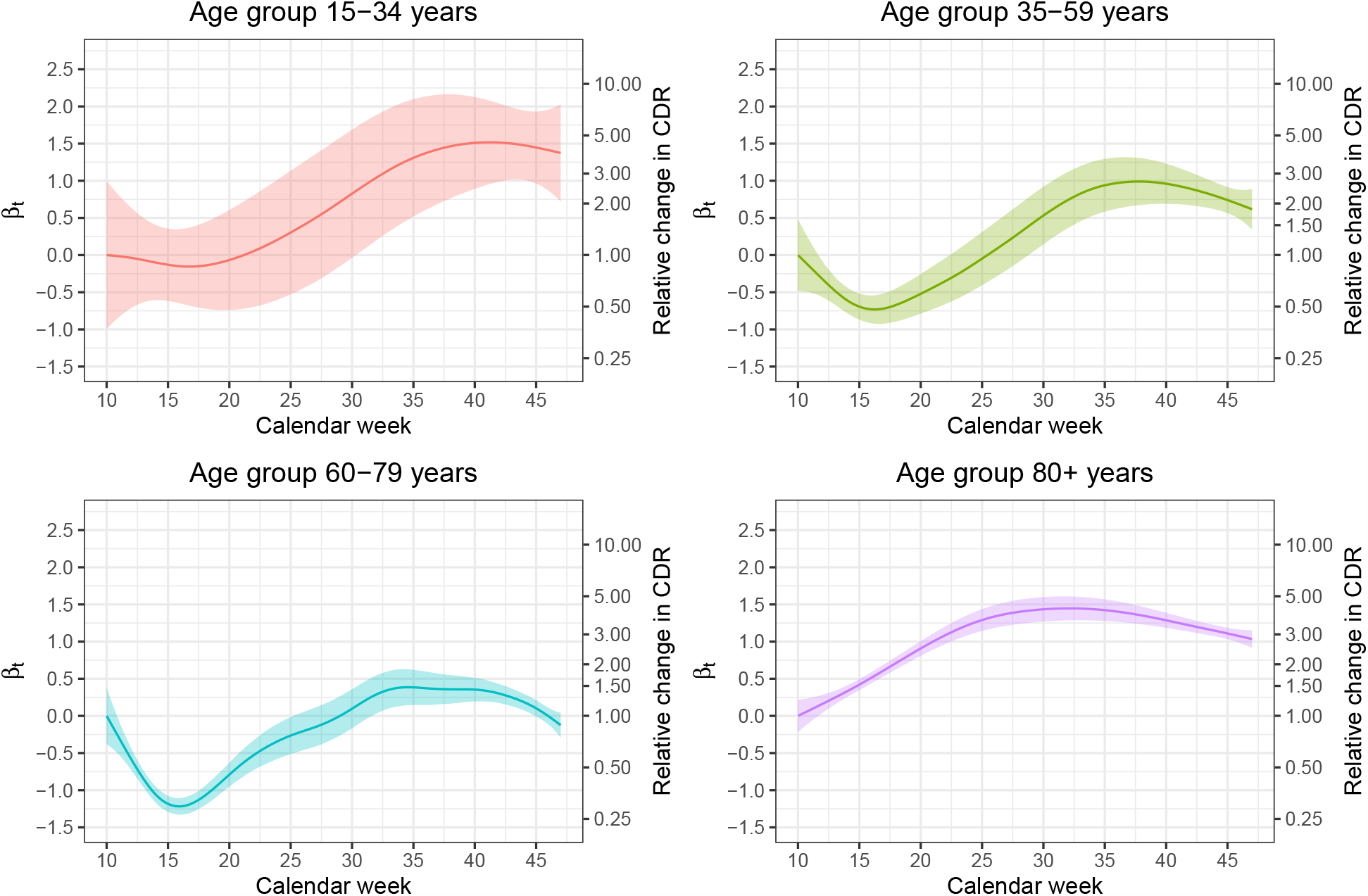
Dynamics in the case-detection ratio for different age groups: The normalized time-varying coefficients *β*_*t*_. The function values on the exp-scale (right y-axes) are the relative change in the case-detection ratio (CDR) with respect to calendar week 10.

## 5. Discussion

Raw reported case numbers and measures derived from them, such as the case fatality ratio, are prone to changes in testing strategies and test capacities, which also influence the case detection ratio. Therefore, a comparison of raw case numbers over time has to be interpreted with care. The case fatality ratio, calculated from the raw number of reported deaths related to COVID-19 divided by reported cases, is additionally impaired by the fact that deaths occur with a time delay after registration, i.e. that deaths registered today correspond to infections which have been reported up to several weeks ago. Our method allows to uncover relative changes in the case detection ratio over different phases of the pandemic. Moreover, we can describe the dynamics in the true number of COVID-19 infections for Germany since March 2020, shedding light on the dark number of undetected cases. The approach is based on publicly available data of registered cases and does not rely on simulations nor on additional survey data. We make use of the fact that, for each fatal outcome, we have the registration date of infection included in the data. This allows us to jointly model the number of registered non-fatal cases and that of fatal infections in a dynamic mixed model, leading to an assessment of the dynamics taking place in real infection numbers. While the plots of raw cases counts in Figure 1 give the impression that the infection incidence was just as high in October as during the peak of the first wave, we could reveal that it was actually still significantly lower, see Figure 2. Based on the available information on the relative change in the case detection ratio over time, we are thus able to compare numbers from the first wave of the pandemic in spring with numbers from the start of the second wave in autumn, adjusting for the difference in the proportion of undetected cases, i.e. the dark number.

For the population aged 80 years and older, the case detection ratio has increased until late summer, when it started to stagnate and then even slightly decreased again. At the same time, in this age group the real number of infections has increased with a concerning slope. As the case detection ratio can be at most 100%, and given that the relative change in this age group was nearly as high as a factor of 4 in CW 33 compared to March, we can conclude that in the beginning of the pandemic the case detection ratio among the population of 80 years and older could not have been more than 25%. Moreover, considering the relative change in the case detection ratio, we can adjust the numbers from the peak in the first wave so that they are comparable e.g. to the numbers in week 41. To exemplify this, note that in week 41 the case detection ratio for age group 80+ was 2.3 times higher as in calendar week 15 during the peak of the first wave. This ratio results from the plot in Figure 3 (bottom right) by taking *β*_*CW* 15_ = 0.4 and *β*_*CW* 41_ = 1.25 and calculating the ratio exp(1.25 − 0.4) = 2.3. In week 41 we had about 18 new infections per week per 100,000 being reported in this age group. In calendar week 15 this number has been 58. However, in week 15 the case detection ratio was much lower as in calendar week 44 and thus we would have seen 2.3 · 58 = 133 cases per 100,000 in this age group 80+ if we had the same CDR in CW 15 as in CW 41.

For the population aged 60-79 years the case detection ratio between the minimum in calendar week 16 and its maximum in calendar week 34 changed by a factor of around five. From this we can deduce that around the peak of the first wave in Germany, at most 20% of the infections were detected, while at least 80% remained unseen. To be able to compare the numbers of the first wave to the numbers in autumn, we apply a similar calculation as above but now for this age group. This results in an estimated number of at least 5 · 16 = 80 cases per 100,000, where only 16 cases per 100,00 have been observed in CW 16.

In age group 35-59 the relative change during the same period was as high as a factor of six, which means that at most one sixth of the infections were detected during week 16. Again, the same calculation shows that the 21 detected infections per 100,000 in week 16 would increase to 6*21=126 cases per 100,000 if we would have had the case detection ratio in week 16 as it was in week 36.

The youngest age group suffers least from severe courses of COVID-19 infections and therefore has the lowest mortality ratios. Consequently, the confidence bands for our estimates in this group are rather wide, while for the oldest age group the estimates are quite precise. We thus refrain from performing similar calculations as above for the age group 15-34, but the general picture is the same as for the other age groups: the case detection ratio has increased from the first wave to the second wave of the pandemic.

A general question in the pandemic is whether large scale testing leads to a high case detection ratio. Our model also allows to investigate this question. In the appendix 6 we refit our model separately for the two most populous German states, North-Rhine-Westphalia and Bavaria. The two states implemented different testing strategies over the summer months. While in Bavaria public test stations were opened in summer, in particular at the borders on the motorways, such fine screening of holiday-returnees was not pursued in North-Rhine-Westfalia. Our model allows to assess and in particular quantify how such different testing strategies lead to different case detection ratios in these two regions. The results show that the Bavarian testing strategy succeeded in reducing the dark figure for the intermediate age groups 35-59 years and 60-79 years when compared to North-Rhine-Westfalia. For the age group 80+ years the course of the case detection ratio is instead practically identical in these two federal states. However, this is not surprising since the additional tests aimed on detecting infected returnees from their holidays and the age group 80+ is the least mobile age group.

## 6. Limitations

A general limitation of our approach is that it suffers from an identifiability issue and hence does not allow to derive absolute numbers for the case detection ratio. However, one may combine our results with findings from seroepidemiological studies. These studies aim to assess on the prevalence of COVID-19 in the general population by screening a representative sample. A list of current seroepidemiological studies in Germany is provided by the RKI (Robert-Koch-Institute, 2020). While these studies provide important information on the current situation of the spread of the disease, they can only give a snapshot of the instantaneous situation at the time the study was conducted. With the knowledge on the dynamics in the infection numbers from our approach, the findings of such studies can be used to predict the situation at other time points. As an example, we look at the Prospective Covid-19 Cohort Study Munich (KoCo19, Radon et al., 2020). They report a CDR of about 25%, where the survey was run between May and June 2020 in the city of Munich. From this number we can deduce a case detection ratio for October being about 3 times higher, concerning the 35-59 age group. More precise calculations would require age specific numbers in the study as well as a regional refit of our model. In principle, however, this demonstrates that the combination of seroepidemiological studies and our approach allows to obtain estimates for absolute numbers of the CDR instead of relative comparisons only.

Moreover, our approach assumes that changes in time do occur steadily, i.e. test capacities are slowly increased and test strategies are gradually changed. This assumption seems in general to be plausible. However, it might not hold when a sudden outbreak in a region causes large parts of the population to be tested. An example is the district of Gütersloh, where after a heavy outbreak in a large industrial slaughterhouse hundreds of infections have been detected after screening. On a national level, however, such local events are averaged out and we can expect that case detection ratios change only smoothly in time and not abruptly.

The most critical assumption of our model is that we assume the infection fatality ratio *a* to be constant over time and negligibly small in comparison to the detection ratio of non-fatal cases. To validate this assumption we performed a sensitivity analysis by refitting the model based on data from May onward only, that is omitting the first wave of the pandemic. The results are provided in the appendix. The picture of the dynamics does not notably change for this refit. To reflect age differences, we have performed all analyses stratified by age group. This age categorisation is rather broad. However, this is a consequence of both the structure of the data and the need for enough cases in a cohort to obtain substantive inference from our model results. In the end, the assumption of a constant fatality ratio also crucially depends on the capacity of the health care system. If this is exceeded, the best possible care for COVID-19 patients is no longer guaranteed. However, the German health system was far from being overburdened during the first wave of the pandemic, and still is.

Finally, it should be noted that our approach does not correct for false positives. Possible effects of misclassification based on the number of tests and assumed specificity ratio are shown by Guenther et al. (2020), which suggests that the real number of infections might even be slightly overestimated from summer onwards.

In conclusion our approach allowed us to show that the true number of infection in Germany in October was considerably lower as during the peak of the first wave, where only a small fraction of COVID-19 infections were detected. By assessing the dynamics in the case detection rate and estimating relative changes we can adjust the case counts reported at different time points so that they become comparable. The analysis of the dynamics in the true infection figures enables a clearer picture of the course of the pandemic.

## Data Availability

All data used for this study are openly accessible.

https://www.arcgis.com/home/item.html?id=f10774f1c63e40168479a1feb6c7ca74

## Appendix

### Sensitivity Analysis

The analysis that we have conducted above has shown that, in Germany, the first COVID-19 wave has peaked around calendar week 14. This became noticeable through higher infection numbers (Figure 2). At the same time we see an estimated local minimum in the case detection ratio for all age groups but the eldest one (Figure 3). To assess the robustness of our approach we now repeat the analysis omitting the data of the first wave and beginning with the first week of May, i.e. calendar week 18. At this time point the first COVID-19 wave had already flattened considerably. Note again that we fit a separate model for every age group. Figure 4 (Figure 5) shows the previous Figure 2 (Figure 3) overlaid by the fitted *V*_*t*_ (*β*_*t*_) as dashed lines when only fitting the model with data from calendar week 18 onward. With very few exceptions the dashed lines lies within the 95% confidence limits of the original smooth functions. Therefore, we can conclude that the assumption of a time-constant age specific IFR is not violated on Germany.

**Figure 4:**
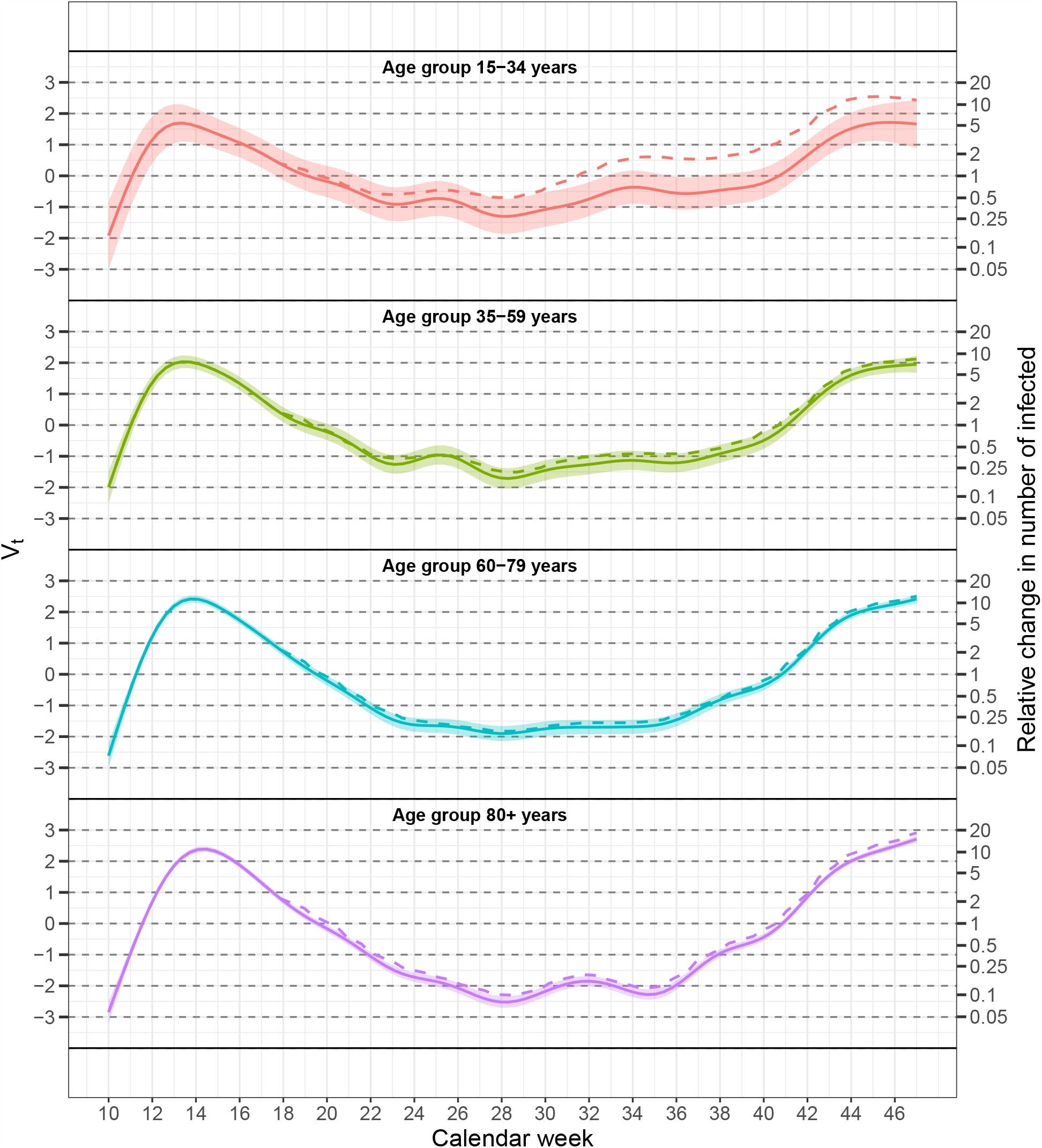
Dynamics of the true infection numbers on the log-scale for different age groups: The smooth random effects *V*_*t*_. Solid lines including 95% confidence bands: Model includes all data; dashed lines: Model included data from calendar week 18 onwards.

**Figure 5:**
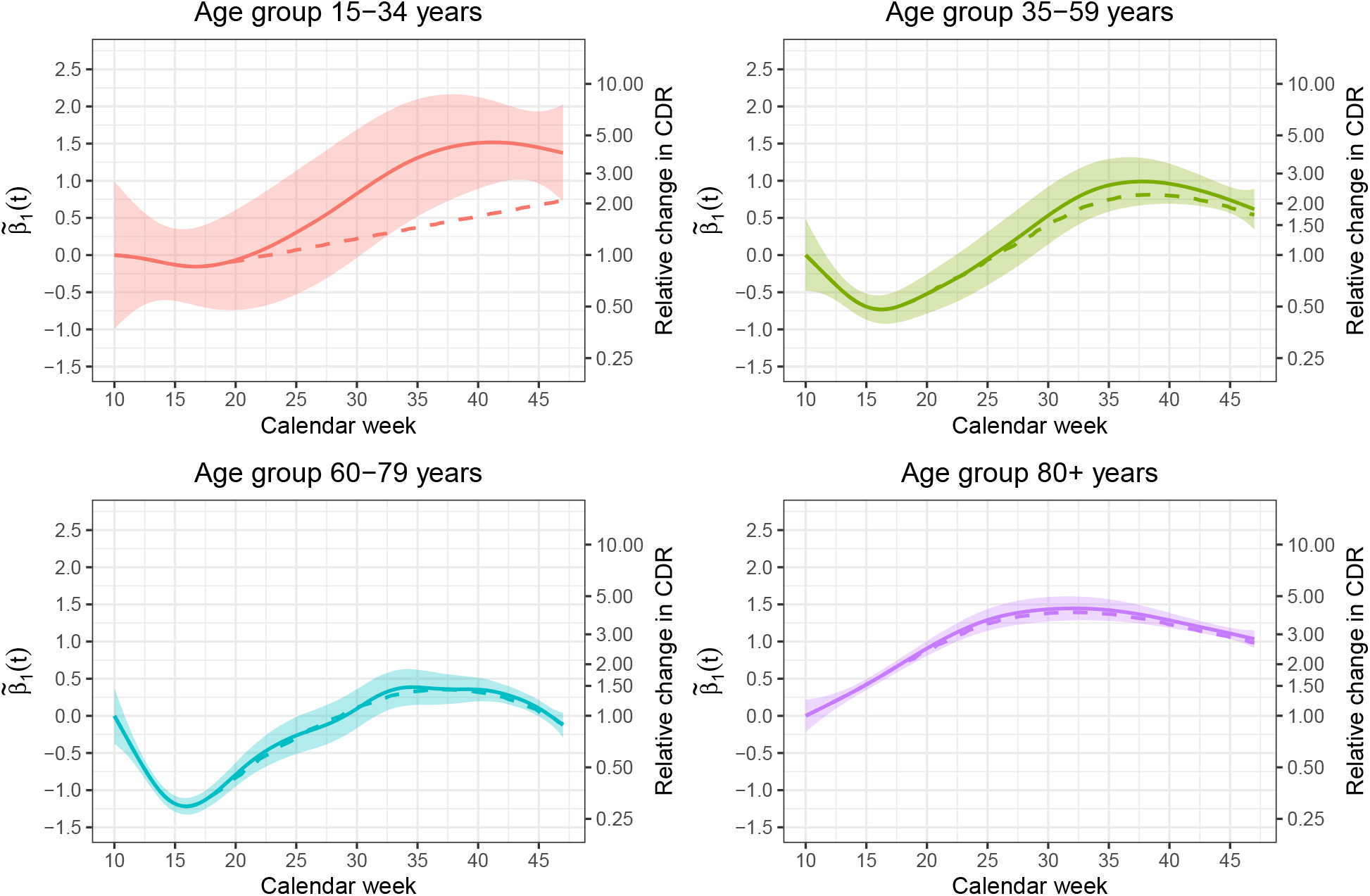
Dynamics in the case-detection ratio for different age groups: The normalized time-varying coefficients *β*_*t*_. The function values on the exp-scale (right y-axes) are the relative change in the case-detection ratio (CDR) with respect to calendar week 10. Solid lines including 95% confidence bands: Model includes all data; dashed lines: Model includes data from calendar week 18 onward.

### Region Specific Analysis

We now look at the results when aggregating the data on the federal state level, and apply our model to data from Bavaria and North-Rhine-Westphalia. These two German federal states are the most populous ones, and have experienced the highest numbers of registered infections. Here, we set again *t* = 1 at calendar week 10 and disregard the age group 15-34 years due to very low fatal infection numbers on the federal state level. Figure 6 shows the random smooth functions *V*_*t*_ on the relative log-scale. While the shape is similar, the scale of *V*_*t*_ is larger for Bavaria than for North-Rhine-Westphalia. Thus, relatively to the early stages of the pandemic, the numbers of infections have dropped more strongly in Bavaria when compared North-Rhine-Westphalia. More fundamentally, Figure 7 shows pronounced differences in the course of the case detection ratio for the intermediate age groups 35-59 years and 60-79 years. That is, the increase in case detection ratio towards the summer months with respect to the beginning of the pandemic is much larger in Bavaria than in North-Rhine-Westphalia. This gives indication that different testing strategies lead to different case detection ratios, a difference which can now be quantified. Note that Bavaria has set up a large number of COVID-19 testing stations and implemented general screening at the borders to Austria, in particular for people returning from the holidays, which was not pursued in North-Rhine-Westfalia.

**Figure 6:**
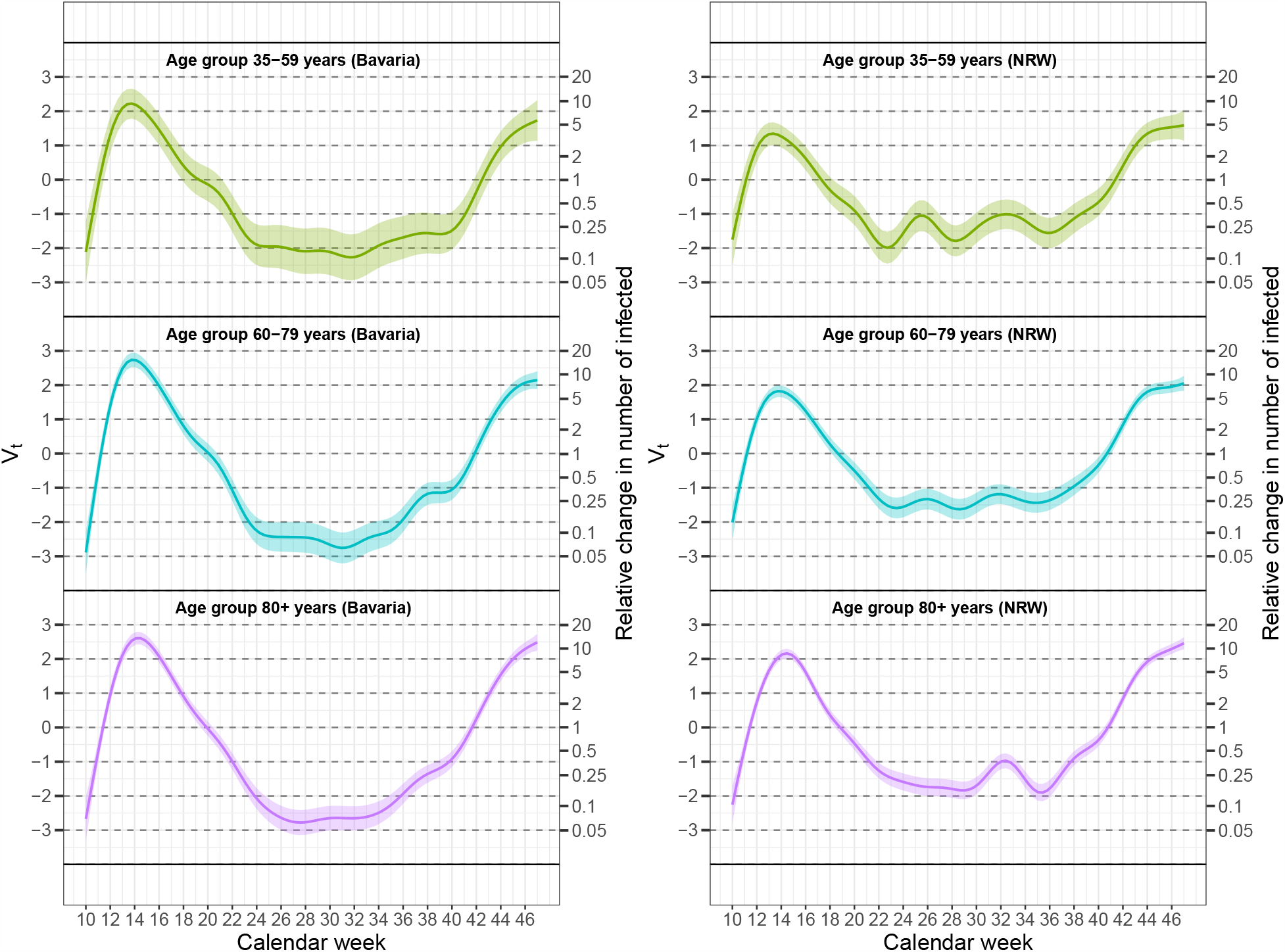
Dynamics of the true infection numbers on the log-scale for Bavaria (left panel) and North-Rhine-Westphalia (right panel): The smooth random effects *V*_*t*_. The shaded areas represent 95% confidence bands.

**Figure 7:**
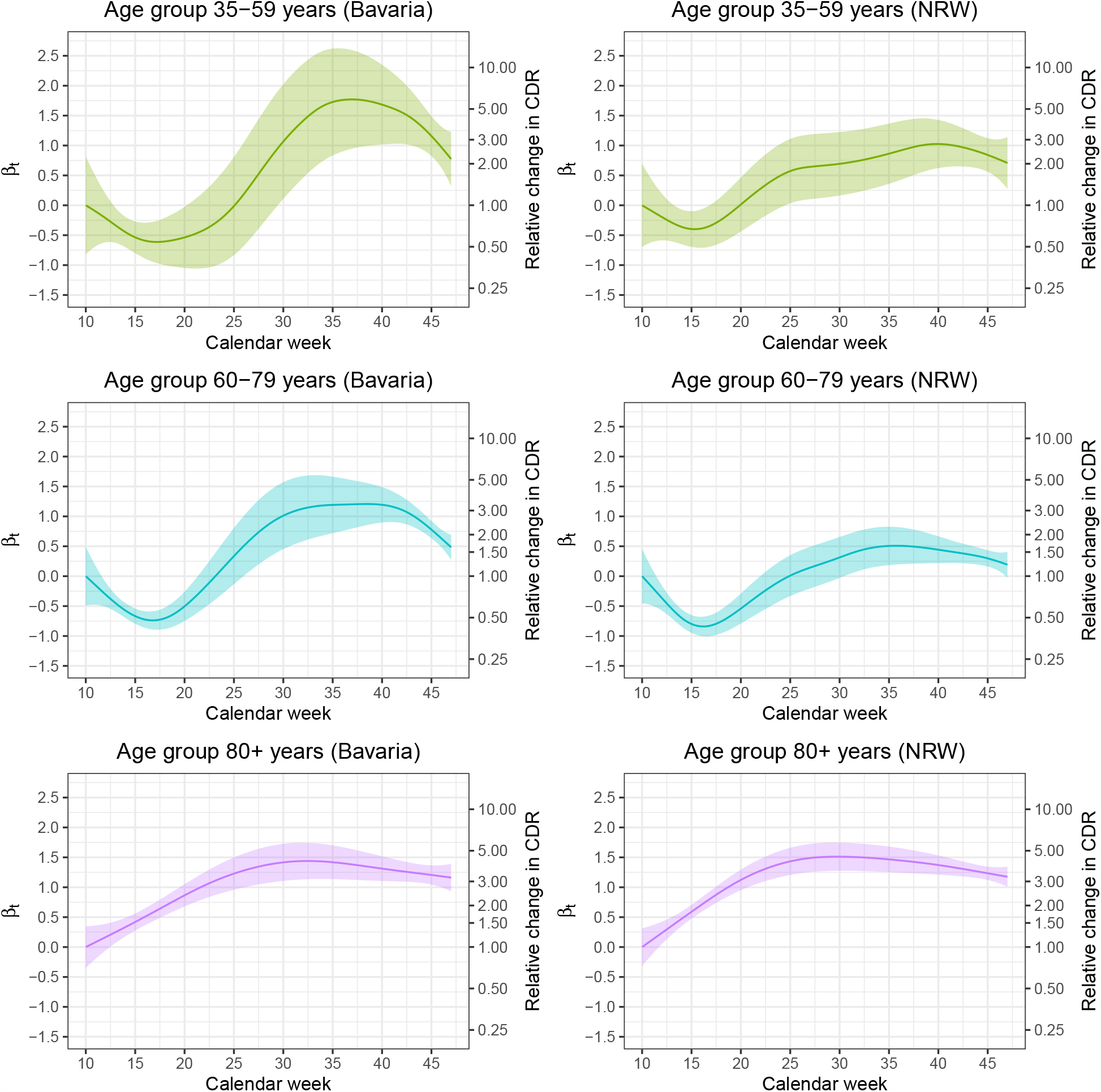
Dynamics in the case-detection ratio for Bavaria (left panels) and North-Rhine-Westphalia (right panels): The normalized time-varying coefficients *β*_*t*_. The function values on the exp-scale (right y-axes) are the relative change in the case-detection ratio (CDR) with respect to calendar week 10.

